# Effect of levodopa/carbidopa on the progression of Machado-Joseph disease /spinocerebellar ataxia type 3 (MJD/SCA3)

**DOI:** 10.1101/2024.11.28.24318145

**Authors:** Mafalda Raposo, Zsuzsa Sárkány, Joana Damásio, CRC-SCA Consortium, Manuela Lima, Sandra Macedo-Ribeiro, Pedro M. Martins

## Abstract

**Background and Objectives:** In Machado-Joseph disease or spinocerebellar ataxia type 3 (MJD/SCA3), ataxin-3 accumulates as neuronal nuclear inclusions in specific regions of the brain such as the substantia nigra and the dentate cerebellar nucleus. Ataxin-3 aggregation is strongly inhibited by dopamine, a neurotransmitter whose brain levels can be increased through the use of medication containing levodopa/carbidopa (LA/CA). Here we perform a retrospective study to determine whether exposition to LA/CA is associated with a decreased progression of MJD/SCA3.

**Methods:** We assessed the natural history of individuals with MJD/SCA3 and also SCA1, 2, 6, 7, 8, and 10 enrolled by the Clinical Research Consortium for Spinocerebellar Ataxias (CRC-SCA). Our initial analysis focused on individuals with all spinocerebellar ataxias (SCAs) whose ataxia progression was assessed by the Scale for the Assessment and Rating of Ataxia (SARA) between 2010 and 2023. To account for important covariates affecting the time evolution of SARA scores, our study was then limited to MJD/SCA3 cases with genetic and age information available. A linear mixed model was used to determine the effects of LA/CA and other anti-parkinsonian drugs at 6 and 13 years of MJD/SCA3 progression.

**Results:** Among the group of 716 individuals with SCAs with monitored SARA scores, 58 (8.1%) had taken LA/CA formulations, whereas 26 (3.6%) had taken other anti-parkinsonian drugs, mainly dopamine receptor agonists. A non-parametric analysis suggested a positive effect of LA/CA on the probability of major disease decline during the initial 6 years of patient monitoring. A multivariate analysis of 262 MJD/SCA3 cases (of which 30 were taken LA/CA) showed statistically significant effects in favor of LA/CA exposition: after adjusting for age and number of CAG repeats in the expanded allele, the yearly SARA response demonstrated a positive effect at 6 years (linear mixed model coefficient **β = −0.651** [95% confidence interval **−1.141 to −0.161**], **P = 0.009**) and 13 years (**β = −0.274** [**−0.547** to **−0.002**], **P = 0.048**).

**Conclusions:** The use of LA/CA to treat non-cerebellar symptoms had a beneficial effect on the SARA score progression in MJD/SCA3. This observation supports the hypothesis that restoring the dopamine levels in the brains of MJD/SCA3 patients might slow down disease progression. Our findings warrant further investigation on whether individuals with MJD/SCA3, even in the absence of Parkinsonian symptoms should also be treated with LA/CA and at what doses.

## Introduction

Machado-Joseph disease or Spinocerebellar ataxia type 3 (MJD/SCA3) belongs to the group of spinocerebellar ataxias (SCAs), which are autosomal dominantly inherited disorders clinically characterized by a progressive loss of balance and coordination, slurred speech and (generally) mid-adulthood onset.^1^ MJD/SCA3 and also SCA 1, 2, 6, 7, 17 and dentatorubral-pallidoluysian atrophy (DRPLA) additionally integrate the group of polyglutamine (polyQ) diseases that currently comprises nine neurodegenerative disorders, including Huntington’s disease. PolyQ diseases are caused by CAG repeat expansions in the coding regions of specific genes that are translated into abnormally long polyQ tracts in disease-characteristic proteins.^2^ The abnormally expanded protein in MJD/SCA3, ataxin-3, aggregates into neuronal nuclear inclusions (NNIs) that also accumulate ubiquitin, ubiquitin-like proteins, proteasome subunits, transcription factors, heat shock proteins and other polyQ proteins.^3^ NNIs are found in affected regions of the MJD/SCA3 brain, such as the cerebellum, substantia nigra, pons and medulla oblongata, but also in areas unaffected by neurodegeneration.^3–5^

While the exact role of ataxin-3 aggregation in MJD/SCA3 remains unclear, the enzymatic activity conferred by the Josephin domain of ataxin-3 is lost upon aggregation.^6^ Furthermore, critical machinery for protein quality control is sequestered during the formation of NNIs.^2^ As with other polyQ proteins, the propensity of ataxin-3 to accumulate into neuronal inclusions correlates well with the length of the expanded polyQ tract; in vitro studies, however, demonstrate that the Josephin domain is crucial for the initial step of protofibril formation, while the expanded polyQ region is decisive in the subsequent step of mature fibril formation.^7^ Non-expanded Ataxin-3 has an intrinsic propensity to aggregate in a polyQ-independent manner.^6–8^ A possible therapeutic strategy for MJD/SCA3 and other polyQ diseases aims at identifying aggregation inhibitors that prevent the occurrence of the toxic gain-of-function associated with protein deposition and the loss-of-function that results from protein sequestration into cellular inclusions.^9,10^ Some of the current clinical trials for MJD/SCA3 prevent protein aggregation by using the autophagy inducer trehalose or allele-specific antisense oligonucleotides (ASOs) that reduce the levels of polyQ expanded and normal ataxin-3, or that bind to expanded CAG repeats in SCA1, MJD/SCA3 and Huntington’s disease.^11^

Novel aggregation inhibitors have been discovered using miniaturized versions of the spectroscopic methods and biophysical assays typically employed to characterize the molecular events in protein self-assembly.^12,13^ Dopamine, for example, was identified as a potent inhibitor of the early steps of secondary nucleation of expanded and non-expanded ataxin-3, as well as of the later steps of fibril maturation of expanded ataxin-3.^14^ Given that dopamine neuronal activity is significantly affected both in patients and asymptomatic mutation carriers of MJD/SCA3,^15^ anti-parkinsonian drugs were hypothesized as repurposing alternatives to prevent ataxin-3 aggregation via the restoration of the dopamine homeostasis.^14^ The authors confirmed that treatment with the dopamine precursor levodopa noticeably improved motor dysfunction in a *C. elegans* model of MJD/SCA3 that expresses full-length pathogenic ataxin-3 in all neurons.^14^ Levodopa remains the standard drug in the treatment of Parkinson’s disease (PD) symptoms. It is available in different formulations combined with carbidopa, a peripherally-acting decarboxylase inhibitor that prevents the systemic conversion of levodopa into dopamine thereby increasing levodopa’s bioavailability in the brain.^16^ Relative to alternatives such as dopamine agonists and monoamine oxidase type B inhibitors, levodopa is associated with greater motor improvement and tolerability but also with higher frequency of dyskinesia.^17^

According to the age at onset and presence of extrapyramidal, pyramidal or neuropathy in addition to a cerebellar syndrome, MJD/SCA3 is classically divided into 3 subtypes to which a fourth subtype characterized by levodopa-responsive Parkinsonism was added afterwards.^18,19^ Moreover, dystonia and restless leg syndrome are among the levodopa-responsive symptoms that have been sporadically reported in MJD/SCA3.^20–22^ In a study involving 75 symptomatic MJD/SCA3 patients, 21 manifested dystonia and 14 completed a 2-month trial on levodopa (600 mg/day), with objective improvements being observed in at least three subjects (21%).^23^ Although no curative treatment is currently available for MJD/SCA3, symptomatic treatments for depression, sleep disorders, Parkinsonism, dystonia, cramps, and pain have been contributing to improve the quality of life of MJD/SCA3 patients.^24^

Longitudinal progression data of ataxia severity has been collected by the Clinical Research Consortium for Spinocerebellar Ataxias (CRC-SCA), which enrolls patients with polyQ SCAs (SCA1, 2, 3, 6 and 7) and other repeat expansion SCAs (SCA8 and 10) from all disease stages.^25,26^ We compared MJD/SCA3 progression in participants previously exposed to levodopa/carbidopa (LA/CA) medication and LA/CA-naive. Interestingly, a favorable effect is identified for LA/CA but not for other anti-parkinsonian drugs at 6 and 13 years since the first clinical evaluation. The advantage of dopamine-restoring therapies (levodopa-based) over dopamine-replacement therapies (such as dopamine agonists) strengthens the hypothesis that dopamine has a disease-modifying effect on MJD/SCA3.

## Methods

### Participants

Our study is based on the CRC-SCA cohort of subjects with SCA1, 2, 3, 6, 7, 8 and 10 which were voluntarily recruited from 12 centers in the US.^26^ The CRC-SCA dataset we assessed contained anonymized information collected from 2010 to 2023 on ataxia severity, CAG repeat expansion, concomitant medication, demographics and comorbidities. The patients seen in ataxia clinics were either self-referred or referred community physicians, local support groups as well as the National Ataxia Foundation. The uniform study protocol was approved by local institutional review boards, and written informed consent was obtained from each participant. The inclusion criteria included (1) diagnosis of the spinocerebellar ataxias in the study subject or his/her affected family members, (2) presence of ataxia, (3) willingness to participate in the study, and (4) the age of 6 years and older. The exclusion criteria were (1) known recessive, X-linked or mitochondrial ataxias, 2) concomitant disorder(s) that affect the Scale for the Assessment and Rating of Ataxia (SARA) and other ataxia measures, or 3) exclusion of the spinocerebellar ataxias by DNA testing.

### Genetic and clinical evaluation

After enrollment in the CRC-SCA study, all participants were asked to provide blood samples for SCA genotyping (non-mandatory).^26^ Information about CAG repeat expansion from samples of 441 participants was obtained via fragment analysis in Dr. Stefan Pulst’s laboratory as described previously.^27^ For participants whose blood samples were unavailable in the research lab, CAG repeat lengths were determined by the commercial laboratories Ataxia severity was evaluated by trained ataxia experts who recorded SARA scores for each patient over different time points, ranging from a single baseline visit up to 16 years of follow-up visits organized by 6-month intervals. SARA scores sum up different rated items, namely gait, stance, sitting, speech, finger chase, nose-finger test, fast alternating hand movements and heel-shin slide, to yield a value between 0 (no ataxia) to 40 (most severe ataxia).^28^

### Definition of Cases and Controls

Our study comprised two non-parametric analyses of the population with SCA1, 2, 3, 6, 7, 8 or 10, and one multivariate analysis of the population with MJD/SCA3 (Figure 1). We selected 716 participants in the CRC-SCA cohort who had documented medication history and SARA scores and who were not considered early-symptomatic. Patients with monitored SARA scores who showed baseline values lower than 8 and a difference between the last SARA score and the baseline lower than 3 were considered early-symptomatic. For the non-parametric Kaplan-Meier analyses, 293 participants who had a single SARA score evaluation were excluded. For the multivariate analysis, only participants with MJD/SCA3 were selected (570 excluded out of 716). In the first non-parametric analysis, controls and cases were defined from the medication history of each participant, namely according to the absence (controls) or existence (cases) of reported use of LA/CA medication. This criterion comprises current or past prescriptions of clinical drugs whose names include the words “levodopa”, “carbidopa”, “sinemet”, “sinomet”, “stalevo”, “dopar” or “larodopa”. For the second non-parametric analysis, the population of 382 participants without recorded LA/CA use was further divided into those who did (cases) or did not (controls) take any of the following anti-parkinsonian drugs: “pramipexole”, “ropinirole”, “requip”, “neupro”, “bromocriptine”,“pergolide”, “cabergoline”, “tolcapone”, “rotigotine”, “entacapone”, “rasagiline”, “mirapex”, or “artane”. From the 146 participants included in the multivariate analysis, 122 controls and 24 cases were selected according to the absence (controls) or existence (cases) of reported use of the LA/CA medication defined above. In addition, a larger group of 155 participants (131 controls and 24 cases) that includes early-symptomatic MJD/SCA3 gene carriers was analyzed. The group with 146 participants was further characterized in terms of reported used of (i) riluzole or troriluzole, and (ii) coenzyme Q10 (searched words: “coenzyme q10”, “co-enzyme Q10”, “coq10”, “co q10”, “co-q10”).

**Figure 1.**
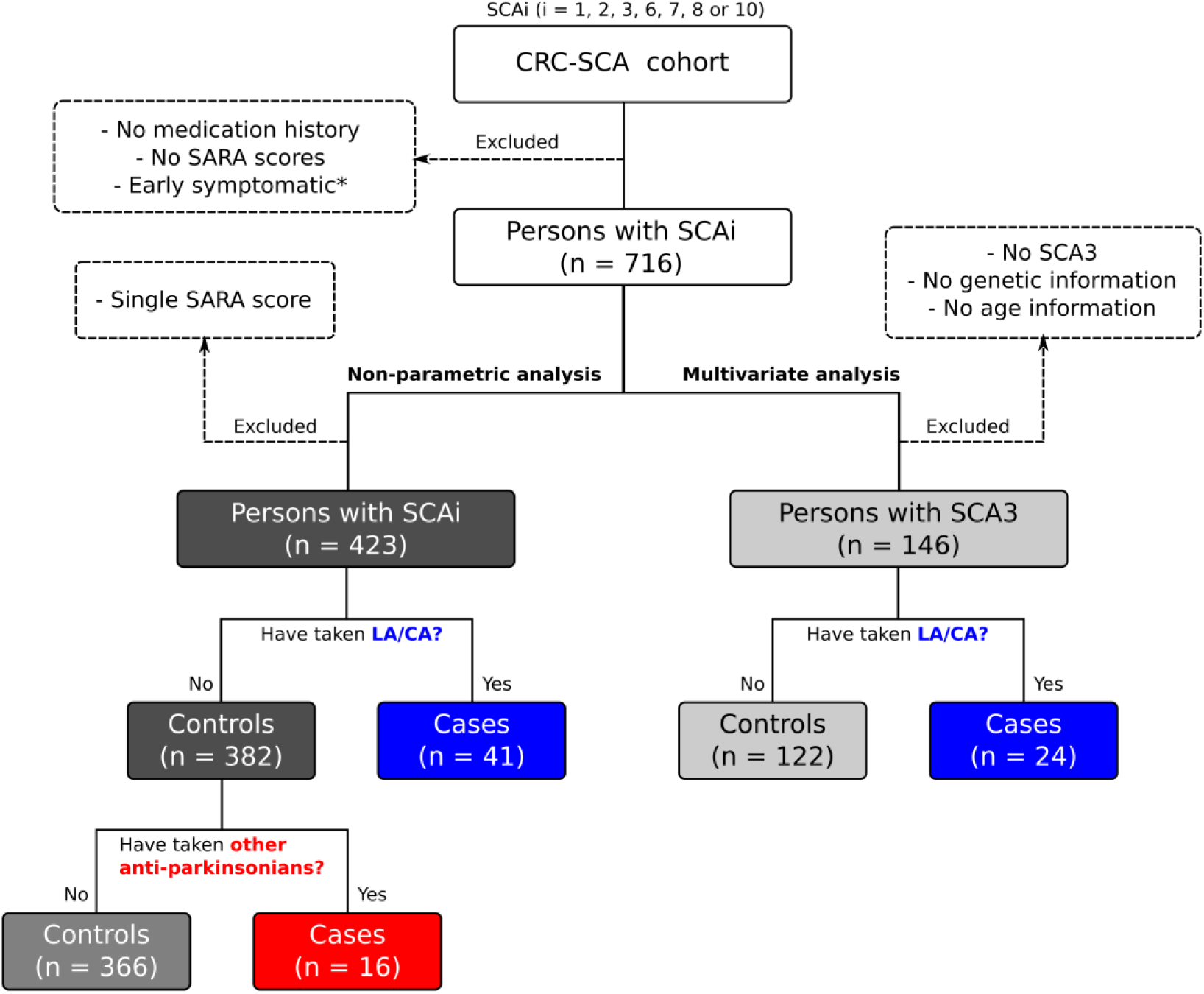
Identification of SCA cases and controls. *Early symptomatic: cases that, simultaneously, have a baseline SARA score lower than 8 and a difference between the last SARA score and the baseline lower than 3.

Statistical analysis. Comparisons of means between groups were performed with a two-sided, unpaired t-test using GraphPad Prism 9. The time from the baseline to the observation of a major disease decline was assessed by the Kaplan-Meier method, where the event of interest corresponds to a SARA score increase larger than 7 points. Kaplan-Meier curves were plotted to compare the probability of event survival for SCA patients with (cases) and without (controls) reported use of LA/CA and other anti-parkinsonian drugs. These non-parametric analyses do not account for confounding factors such as the type of spinocerebellar ataxia, age, and the length of expanded allele repeat. To control for covariates, we focused on the most common SCA (MJD/SCA3) and selected cases and controls for which genetic and age information was available (Figure 1). A linear mixed-effects (LME) model regression was performed to determine whether there was a significant change in the time evolution of SARA scores upon exposition to LA/CA medication. The adopted Wilkinson notation for the SARA score response was the following: **SARA∼TREATMENT + TIME + TREATMENT: TIME + AGE + ALLELE + (1|PATIENT)**, where **TREATMENT** denotes whether the patient was exposed to LA/CA or not, **TIME** represents the number of years since the baseline visit, **TREATMENT: TIME** is the interaction term, **AGE** and **ALLELE** are additional covariates computed in years and number length of expanded allele, respectively, and **1|PATIENT** denotes random intercepts assigned to individual patients. The model was regressed using the *fitlme* function of MATLAB R2023a (MathWorks, Natick, MA) to determine LME coefficients (**β**) and the associated P values. The LME model fitted to the longitudinal SARA increase at 6 years was used to predict the evolution of SARA scores for a 56-year-old patient with an expanded allele length of 56. P values less than 0.05 were considered significant.

## Results

### Use of LA/CA and other anti-parkinsonian drugs by individuals with spinocerebellar ataxias

By assessing the CRC-SCA records of concomitant medication use, we identified 58 SCA patients (8.1%) who were prescribed with levodopa in formulations that contained carbidopa and, occasionally also, entacapone (Table 1). Other anti-parkinsonian drugs, such as dopamine agonists, have also been used together with, or as an alternative to LA/CA. In the present study, we refer to “other anti-parkinsonian drugs” as corresponding to non-LA/CA anti-parkinsonians prescribed to SCA patients who have no track record of LA/CA use (Figure 1). Overall, 26 cases were identified meeting this criterion (Table 1). Exposition to LA/CA was verified in all spinocerebellar ataxias except SCA7. The larger populations of MJD/SCA3 and SCA2 patients were also those associated with a more common use of LA/CA medication (11.4% among 264 MJD/SCA3 patients, and 9.4% among 159 SCA2 patients). Cases with a history of taking anti-parkinsonian drugs had, in this sample, larger CAG repeats in the expanded allele than the control groups (Tables 1 and S2).

**Table 1.** Characteristics of the 716 participants with available medication history and quantified disease severity.

|  | LA/CA |  | OTHER ANTI-PARKINSONIANS |  |
| --- | --- | --- | --- | --- |
|  | Controls | Cases | Controls | Cases |
| N | 658 | 58 | 632 | 26 |
| SCA1 | 108 | 4 | 103 | 5 |
| SCA2 | 144 | 15 | 138 | 6 |
| MJD/SCA3 | 234 | 30 | 221 | 13 |
| SCA6 | 123 | 6 | 122 | 1 |
| SCA7 | 14 | 0 | 14 | 0 |
| SCA8 | 27 | 2 | 26 | 1 |
| SCA10 | 8 | 1 | 8 | 0 |
| Female, n (%) | 359 (55) | 31 (53) | 346 (55) | 13 (50) |
| Age (years)* | 52.5 ± 14.1 | 50.6 ± 14.2 | 52.5 ± 14.2 | 54.0 ± 13.0 |
| Expanded allele repeat length* | 50.9 ± 24.0 | 57.3 ± 18.9 | 50.7 ± 24.3 | 56.2 ± 17.8 |

\*\*Mean ± standard deviation (S.D.) values; missing data ignored. Refer to Table S2 for comparisons of means between groups.

Parkinsonism, tremor, dystonia, bradykinesia and restless legs syndrome are among the indications appointed for LA/CA prescription (Table 2). There are, however, many cases of unspecified indications or general references to ataxia management. To ascertain a possible disease-modifying effect of LA/CA drugs, we started by looking for major changes in disease progression among the whole sample of spinocerebellar ataxias, thus harnessing the data collected in a total of 1380 ataxia assessments (Figure 2A). To control for unspecific effects, we also considered the group of patients who were prescribed anti-parkinsonian drugs but not LA/CA (Figure 2B). The subsequent filtering out of cases other than MJD/SCA3 or with missing information produced a sample with 146 MJD/SCA3 patients who had undergone 354 medical visits for ataxia rating and assessment (Figure 2C).

**Table 2.** Symptoms underlying the use of LA/CA formulations in spinocerebellar ataxias.

| Symptoms | n (%) |  |  |  |  |  |
| --- | --- | --- | --- | --- | --- | --- |
|  | SCA1 | SCA2 | MJD/SCA3 | SCA6 | SCA8 | SCA10 |
| Tremor, dystonia, bradykinesia, dyskinesia | 0 (0) | 7 (47) | 3 (10) | 1 (17) | 1 (50) | 0 (0) |
| Parkinsonism | 0 (0) | 0 (0) | 2 (7) | 2 (33) | 0 (0) | 0 (0) |
| Restless legs syndrome | 0 (0) | 1 (7) | 5 (17) | 1 (17) | 0 (0) | 0 (0) |
| Ataxia | 3 (75) | 4 (27) | 5 (17) | 1 (17) | 1 (50) | 1 (100) |
| Other/unspecified | 1 (25) | 3 (20) | 15 (50) | 1 (17) | 0 (0) | 0 (0) |
| Total | 4 (100) | 15 (100) | 30 (100) | 6 (100) | 2 (100) | 1 (100) |

**Figure 2.**
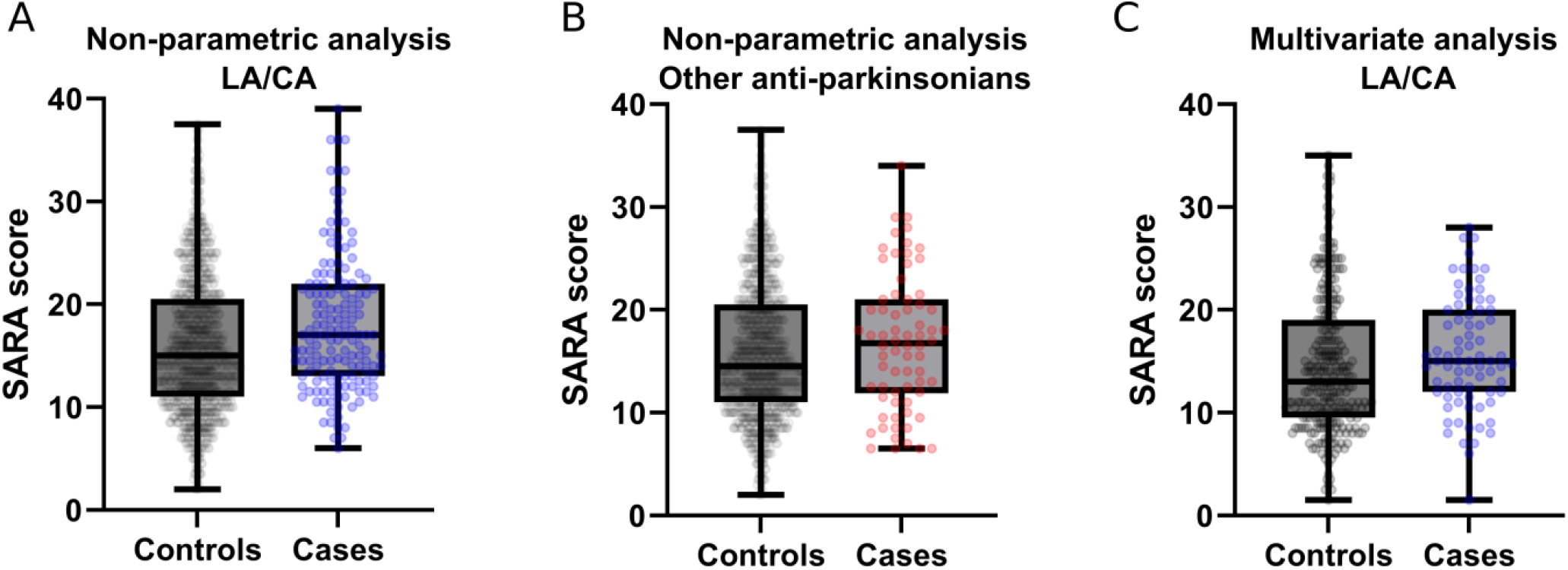
Boxplots of all measured SARA scores. The boxes extend from the 25th to 75th percentiles, the central line is the median and the whiskers represent minimum and maximum values. The symbols are measured data used in our (A and B) non-parametric and (C) multivariate analyses. Number of visits: (A) 1231 controls and 149 LA/CA cases; (B) 1161 controls and 70 cases of other anti-parkinsonian drugs; (C) 278 controls and 76 LA/CA cases.

### Non-parametric analyses

To investigate the progression of SCAs, only participants with multiple SARA records were included (423 out of 716). Of these, 35.9% had a diagnosis of MJD/SCA3, 24.3% of SCA2 and 14.9% of SCA1 (Table 3). Again, exposition to anti-parkinsonian medication was associated with more CAG repeats in the expanded allele. Participants in the LA/CA group are slightly younger than controls, whereas the group receiving other anti-parkinsonian drugs is, on average, 2 years older than the control group.

**Table 3.** Characteristics of the 423 participants selected for the non-parametric analyses.

|  | LA/CA |  | OTHER ANTI-PARKINSONIANS |  |
| --- | --- | --- | --- | --- |
|  | Controls | Cases | Controls | Cases |
| n | 382 | 41 | 366 | 16 |
| >6 years monitoring, n* | 51 | 7 | 49 | 2 |
| SCA1 | 60 | 3 | 59 | 1 |
| SCA2 | 92 | 11 | 88 | 4 |
| MJD/SCA3 | 132 | 20 | 123 | 9 |
| SCA6 | 74 | 5 | 73 | 1 |
| SCA7 | 4 | 0 | 4 | 0 |
| SCA8 | 13 | 2 | 12 | 1 |
| SCA10 | 7 | 0 | 7 | 0 |
| Female, n (%) | 205 (54) | 24 (59) | 196 (54) | 9 (56) |
| Age (years)** | 53.2 ± 14.0 | 52.4 ± 12.5 | 53.1 ± 14.1 | 55.1 ± 12.7 |
| Expanded allele repeat length** | 49.7 ± 23.6 | 56.2 ± 19.8 | 49.4 ± 23.8 | 56.1 ± 19.7 |

\*SARA scores monitored over periods longer than 6 years
\*\*Mean ± S.D. values; missing data ignored. Refer to Table S3 for comparisons of means between groups.

To visualize the overall effects of LA/CA and other anti-parkinsonian medications, we computed the time required for major disease progression events to occur in cases and controls. We defined these events as corresponding to an increase in the SARA score larger than 7 points relative to the initial assessment (Figure 3); subsequently, a threshold difference of 4 points was also tested (Figure S1). Kaplan-Meier plots of event probability vs. time suggest that the LA/CA group had a lower susceptibility to a major disease decline over the 6 years that followed the initial ataxia assessment. This apparent advantage was lost in the post-6-year period (Figure 3A). Conversely, the group of other anti-parkinsonian drugs showed no major differences to controls over the initial 6 years and appeared to slightly improve in the subsequent period (Figure 3B). If the threshold SARA score increase is set to 4 (rather than 7), treatment with LA/CA still compares negatively with controls and treatment with other anti-parkinsonian drugs (Figure S1). Importantly, the groups being compared comprised different ataxias, were not paired in terms of age and genetic burden, and involved a reduced number of participants monitored for periods longer than 6 years (Table 3).

**Figure 3.**
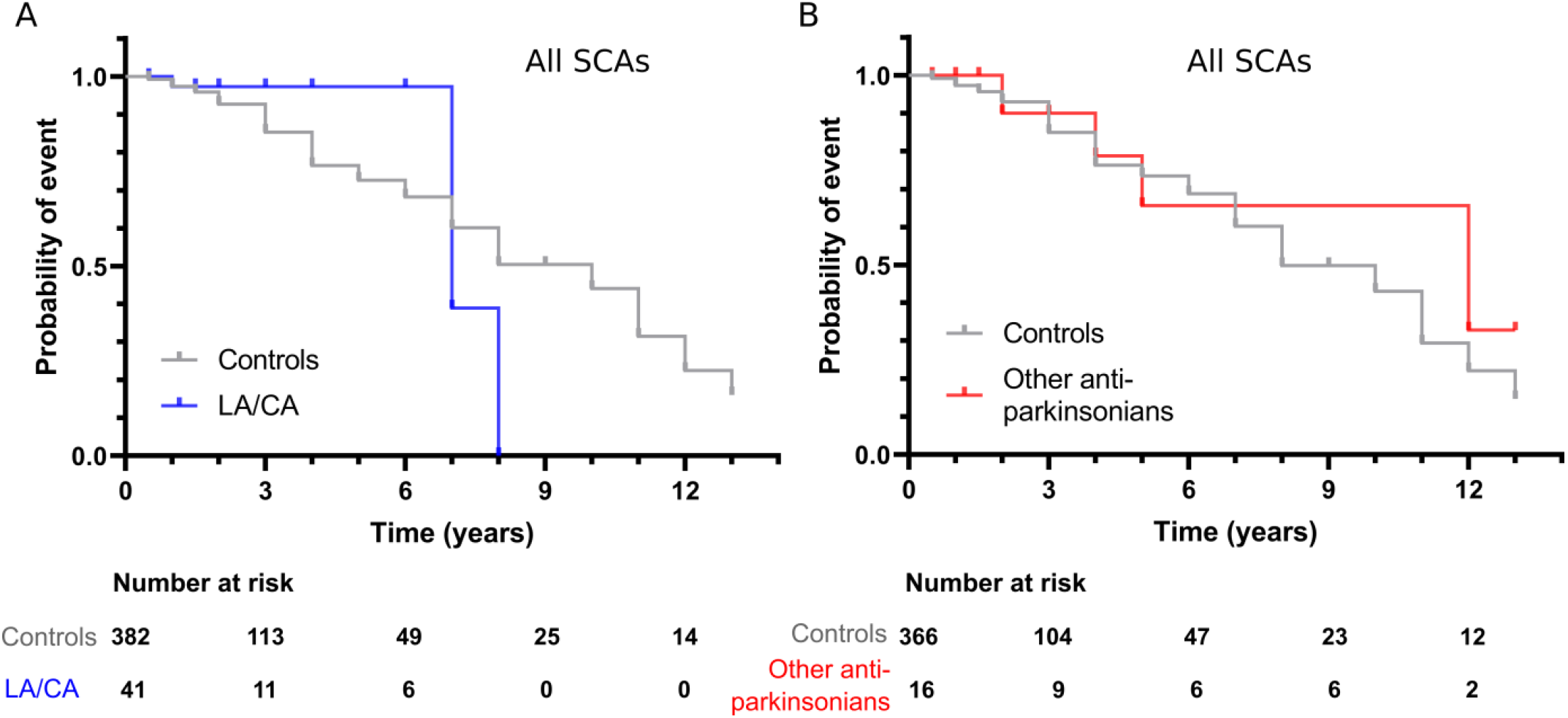
Resistance to major disease decline in spinocerebellar ataxias: the effect of anti-parkinsonian drugs. (A and B) Kaplan-Meier analysis for events of a large SARA score increase (> 7 points) in patients (A) who have previously taken LA/CA compared with controls, and (B) who have previously taken other anti-parkinsonian drugs compared with controls.

### Multivariate analysis

To account for confounding factors affecting disease progression, we centred our analysis on the most represented disease (MJD/SCA3) and on participants with known age and CAG repeat length. A linear mixed model was employed to evaluate the effect of LA/CA exposition adjusted for covariates, thus taking full advantage of a SARA score dataset in which participants with a single medical visit were also included. The number of MJD/SCA3 patients meeting the selection criteria is 148, of which 24 have been prescribed with LA/CA (Table 4).

**Table 4.**
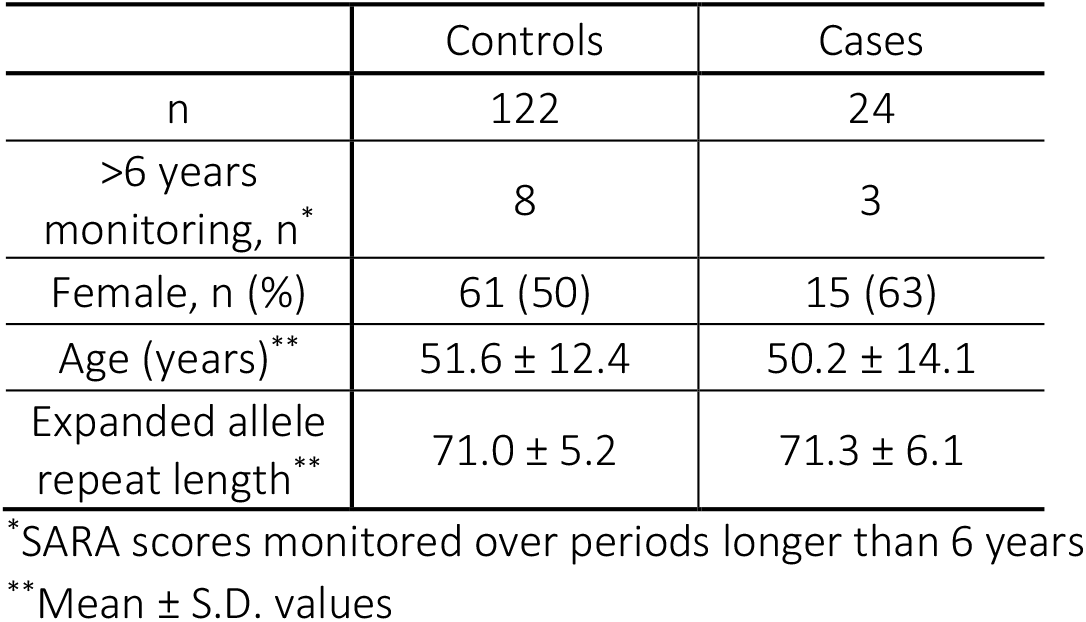
Characteristics of the MJD/SCA3 group selected for the multivariate analysis of the LA/CA effect.

Through multivariate analysis, significant effects of age, time since the first visit, CAG repeat length and LA/CA treatment are identified from the measured data of SARA score progress at 6 and 13 years (Figure 4 and Table 5). As expected, age, time and repeat length were characterized by positive values of the LME coefficient β (Table 5), meaning that SARA scores increased with the covariate magnitude. These effects were confirmed during the analyses of drug exposition to LA/CA but also to non-LA/CA anti-parkinsonian drugs (Tables S4 and S5, and Figure S2) and anti-parkinsonians in general (Tables S6 and S7, and Figure S3). For LA/CA, the interaction term confirmed a statistically significant effect in favour of drug exposition (negative β) both at 6 and 13 years. Such an effect was not observed for other anti-parkinsonian drugs (Table S5). A total of 9 MJD/SCA3 gene carriers (none of which with a history of anti-parkinsonian medication) are here considered early-symptomatic because they cumulatively show a baseline SARA score lower than 8 and a difference between the last SARA score and the baseline lower than 3 (Figure 1). If these cases are additionally considered in the group of controls, the effect of LA/CA becomes weaker (Figure S4 and Tables S8 and S9) but is still significant at 6 years (**P = 0.036**).

**Table 5.** Covariate-adjusted effect of LA/CA exposition on MJD/SCA3 progression. LME coefficients, 95% confidence intervals and P values fitted to the 13-year and 6-year SARA monitoring data.

| 13 YEARS |  |  |  |  |
| --- | --- | --- | --- | --- |
| EFFECT | $\beta$ | Lower | Upper | P value |
| INTERCEPT | -32.89 | -52.40 | -13.4 | 0.001 |
| AGE | 0.23 | 0.14 | 0.3 | <0.001 |
| TIME | 0.99 | 0.81 | 1.2 | <0.001 |
| ALLELE | 0.48 | 0.25 | 0.7 | <0.001 |
| TREATMENT | 2.25 | -0.34 | 4.8 | 0.088 |
| **TIME:**<br>**TREATMENT** | **-0.27** | **-0.55** | **0.0** | **0.048** |
| 6 YEARS |  |  |  |  |
| INTERCEPT | -32.68 | -52.24 | -13.1 | 0.001 |
| AGE | 0.23 | 0.14 | 0.3 | <0.001 |
| TIME | 1.01 | 0.76 | 1.3 | <0.001 |
| ALLELE | 0.48 | 0.25 | 0.7 | <0.001 |
| TREATMENT | 2.46 | -0.15 | 5.1 | 0.064 |
| **TIME:**<br>**TREATMENT** | **-0.65** | **-1.14** | **-0.2** | **0.009** |

**Figure 4.**
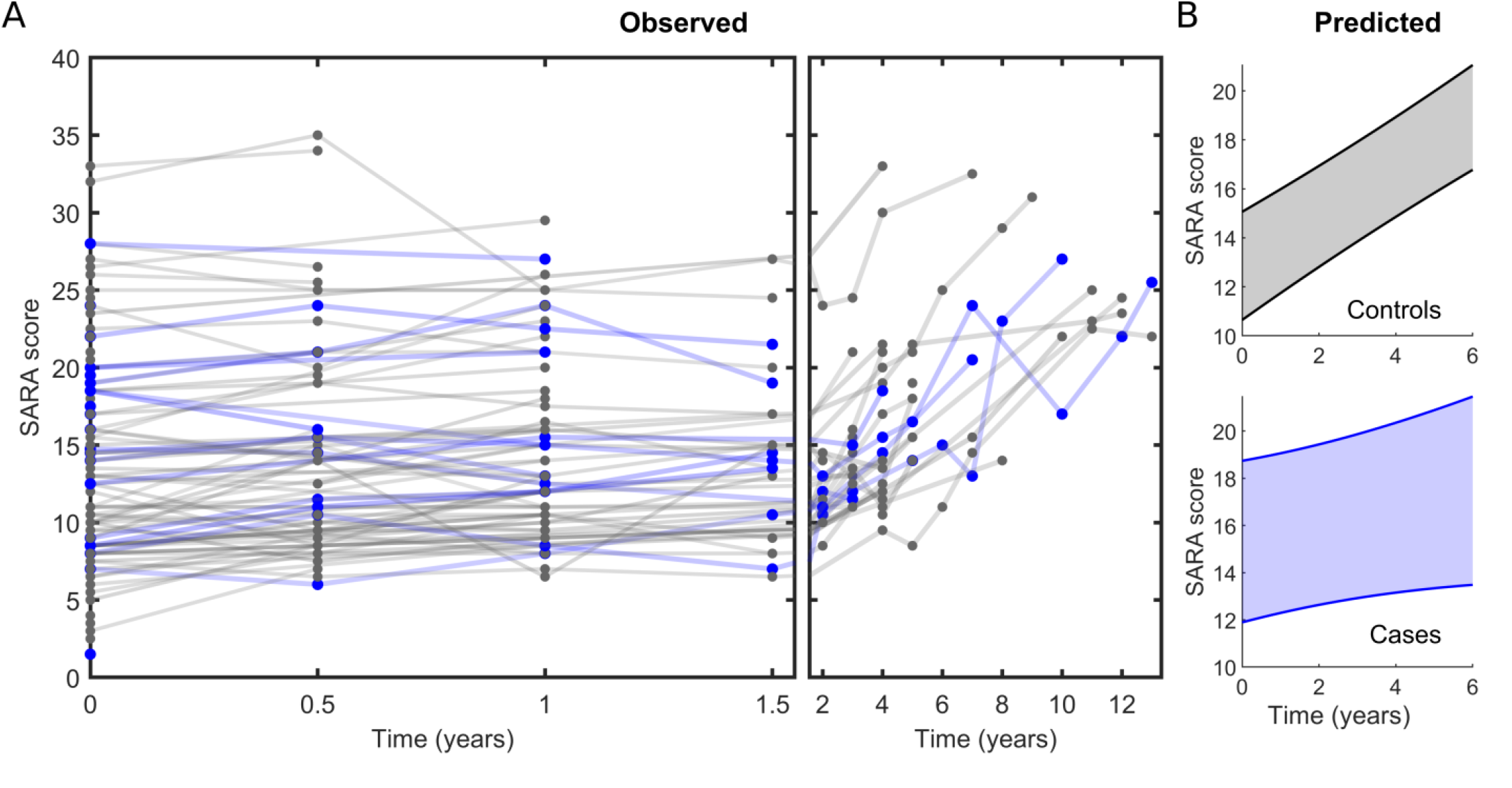
Effect of exposition to LA/CA on MJD/SCA3 evolution. Progress of SARA scores (A) observed in individuals with MJD/SCA3 and (B) predicted by the LME model for (top) controls and (bottom) cases of exposition to LA/CA. (B) 95% confidence intervals for 6-year predictions computed assuming a random MJD/SCA3 patient, 58 years of age, and expanded allele length 70.

The non-parametric analysis of all ataxias suggested a beneficial effect of LA/CA on ataxia decline events over a 6-year interval (Figure 3A); the MJD/SCA3-restricted multivariate analysis confirmed a positive effect of LA/CA exposition on the longitudinal SARA increase over the same time interval. Although the interaction term coefficients denote a weaker influence at 13 years (Table 5), this effect is still statistically significant in the long term. The fitted LME model was used to predict the evolution of SARA scores in LA/CA-exposed and -naive individuals with MJD/SCA3. By considering the 6-year timeframe and average values of age and length of expanded allele, an increase in the SARA score of 2.1 points is predicted for LA/CA exposed patients, while the corresponding variation in controls amounts to 6.0 points (Figure 4B).

## Discussion

Our study has produced two types of results: one, of a more descriptive nature, relies on a non-parametric analysis of all SCAs irrespective of each disease’s peculiarities, participant’s age or genetic burden. The second type is free from confounding factors associated with multiple disease analysis since solely MJD/SCA3 was considered. Moreover, using a linear mixed model we captured and statistically adjusted the expected influence of age and length of expanded allele repeat on the longitudinal variation of SARA scores. Both types of results point to favorable effects associated with the use of LA/CA drug formulations. If all SCA participants are considered, major disease decline is delayed for cases of LA/CA exposition during the first 6 years of disease monitoring but, in the subsequent period, this higher resistance becomes no longer evident (Figure 3A). The multivariate analysis centered on MJD/SCA3 participants with fully characterized profiles showed that disease progression at 6 and 13 years is delayed relatively to controls for LA/CA cases (Figure 4) but not for other anti-parkinsonian drugs (Figure S2). The benefits of LA/CA exposition by MJD/SCA3 patients do not seem to directly arise from an improvement in Parkinsonian symptoms because, first, SARA score does not seem to be influenced by Parkinsonism,^29^ and second, non-LA/CA anti-parkinsonian drugs did not influence the ataxia progression of MJD/SCA3 isolated or considered together with other ataxias. By considering the use of LA/CA and/or other anti-parkinsonians together, a trend in favor of drug exposition is still obtained, although the fitted values of LME coefficients evidence, as expected, a weaker effect than the observed for CA/DA alone (Figure S3 and Tables S6 and S7).

Despite the promising evidence for disease-modifying effects caused by LA/CA medication, the existence of a 6-year threshold after which symptom aggravation becomes prominent in several SCA patients (Figure 3A) is suggestive of loss of protection or levodopa-induced adverse effects. In the groups of other anti-parkinsonian drugs and controls, the events of major disease decline comprise patient-to-patient variability, i.e., they show a gradual trend if represented in Kaplan-Meier plots (Figure 3B). From previous studies in PD, we know that the mean time from PD symptom onset to the development of levodopa-induced motor complications is, in the case of motor fluctuations, 6.5 (S.D. 4.1) years and, in the case of dyskinesias, 6.7 (S.D. 3.1) years.^30^ In addition, there are several case reports of levodopa-induced dyskinesia in spinocerebellar ataxia patients. ^31–35^ We admit that the delayed MJD/SCA3 progression observed at 6 and 13 years is achieved notwithstanding a long-term aggravation of motor symptoms that is disease-independent and could be associated with long-term levodopa. To confirm this hypothesis, further studies are required with better-controlled LA/DA dosage regimens and treatment duration.

How to interpret a disease-modifying effect of LA/CA on MJD/SCA3 and the absence of such an effect when other anti-parkinsonian drugs are used? A possible explanation is that dopamine levels in the affected brain regions are increased by LA/CA but not by dopamine replacement therapies such as dopamine agonists. In the long run, increased dopamine levels may delay the formation of neuronal inclusions characteristic of MJD/SCA3 by a mechanism of inhibition of ataxin-3 aggregation.^14^ On the other hand, long-duration LA/CA leads to dyskinesias and other motor complications that are less frequently observed for other anti-parkinsonian drugs and may indirectly affect cerebellar symptoms. More intricate mechanisms of LA/CA action involving other neurotransmitter pathways besides the dopaminergic are however admissible taking into account the pharmacodynamics of levodopa,^16^ as well as the complexity of MJD/SCA3 pathogenesis.^2^

Translating these findings into new therapies for MJD/SCA3 patients without Parkinsonian symptoms will necessitate randomized clinical trials conducted under more rigorously controlled conditions than the present study. One of the limitations of our study is that it does not account for the possible beneficial effects of concomitant drugs on MJD/SCA3 progression. For example, among the MJD/SCA3 group in Table 4, 3.3% (4/122) of the controls and 12.5% (3/24) of the LA/CA patients have a history of treatments with riluzole or its prodrug troriluzole, which may delay ataxia progression in patients with various types of degenerative ataxia.^36,37^ On the other hand, 2.4% (3/122) of the controls and 12.5% (3/24) of the LA/CA patients in Table 4 have been prescribed coenzyme Q10, which may be associated with a better clinical status in SCA1 and MJD/SCA3.^38^ Defining what should be the appropriate LA/CA dosage is also important for future trials, given the potential adverse effects of long-term levodopa and the specific challenges associated with dopaminergic impairment in MJD/SCA3. Although the nigrostriatal dopaminergic system is affected in both symptomatic and asymptomatic MJD/SCA3 carriers,^15,39,40^ this feature is likely distinct from the observed in PD regarding the rare number of MJD/SCA3 patients that show levodopa-responsive Parkinsonism. Distinguishing between pre-ataxic, early-symptomatic and ataxic patients is also important for the design of new therapies.^41^ Our results suggest that LA/CA treatments will benefit MJD/SCA3 patients already showing aggravating signs of ataxia. Leveraging the power of levodopa to restore dopamine levels while avoiding its adverse effects may be achieved through novel methods of drug delivery that restore brain dopamine in a more physiological manner and without peaks of abnormal neurotransmitter release.^42^ Alternatively, MJD/SCA3-dedicated formulations might be developed containing LA/CA amounts adjusted to correct dopaminergic deficits milder than in PD.

## Data Availability

All data produced in the present study are available upon reasonable request to the authors

## Acknowledgements

This work was funded by National Funds through FCT—Fundação para a Ciência e a Tecnologia, I.P., under the project UIDB/04293/2020 and PTDC/QUICOL/2444/2021. PMM is supported by FCT CEECIND/03750/2017/CP1386/CT0014

(https://doi.org/10.54499/CEECIND/03750/2017/CP1386/CT0014). MR is supported by FCT CEECIND/03018/2018/CP1556/CT0009 (https://doi.org/10.54499/CEECIND/03018/2018/CP1556/CT0009).

CRC-SCA Consortium composition:

1. Tetsuo Ashizawa, MD and Andrew Billnitzer, MD, MPH Houston Methodist Research Institute, Houston, Texas, USA
2. Susan Perlman, MD Department of Neurology, University of California Los Angeles, Los Angeles, CA USA
3. Khalaf Bushara, MD Department of Neurology, University of Minnesota, Minneapolis, MN USA
4. Michael D Geschwind MD, PhD and Cameron Dietiker, MD Department of Neurology, University of California San Francisco, San Francisco, CA USA
5. Christopher M. Gomez, MD, PhD and Mahesh Padmanaban, MD Department of Neurology, University of Chicago, Chicago, IL USA
6. Sheng-Han Kuo, MD Department of Neurology, Columbia University Medical Center, New York, NY USA
7. Puneet Opal, MD, PhD and Rizwan Akhtar, MD, PhD Department of Neurology, Northwestern University, Chicago, IL USA
8. Henry Paulson, MD, PhD, Sharan Srinivasan, MD, PhD, and Amy Ferng, MD Department of Neurology, University of Michigan, Ann Arbor, MI USA
9. Chiadi U. Onyike, MD Department of Psychiatry and Behavioral Sciences, Johns Hopkins University, Baltimore, MD USA
10. Sarah Ying, MD, Liana Rosenthal, MD, PhD, and Ashley Paul, MD Department of Neurology, Johns Hopkins University, Baltimore, MD, USA
11. Jeremy Schmahmann, MD, Christopher Stephen, MD, Anoopum Gupta, MD, PhD, and Chih-Chun Lin, MD, PhD Department of Neurology, Massachusetts General Hospital, Harvard Medical School, Boston, MA USA
12. S.H. Subramony, MD and Matthew Burns, MD, PhD Department of Neurology, University of Florida, Gainesville, FL USA
13. George Wilmot, MD, PhD Department of Neurology, Emory University, Atlanta, GA USA
14. Antoine Duquette, MD, MSc Centre Hospitalier de l’Université de Montréal, University of Montreal, Montreal, QC Canada
15. Theresa Zesiewicz, MD Department of Neurology, University of South Florida, Tampa, FL USA
16. Marie Y. Davis, MD, PhD Department of Neurology, University of Washington, Seattle, WA USA
17. Ali G. Hamedani, MD, MHS and Joaquin Vizcarra Pasapera, MD Department of Neurology, Perelman School of Medicine, University of Pennsylvania, Philadelphia, PA USA
18. Vikram G. Shakkottai, MD, PhD Department of Neurology, University of Texas Southwestern Medical Center, Dallas, TX USA
19. Stefan Pulst, MD Department of Neurology, University of Utah, Salt Lake City, UT USA
20. Lauren Moore, PhD National Ataxia Foundation, Minneapolis, MN USA

