## Supplementary Figures and Tables for "Effect of levodopa/carbidopa on the progression of Machado-Joseph disease /spinocerebellar ataxia type 3 (MJD/SCA3)"

Table S1. List of anti-parkinsonian drugs considered in this study.

| Type | Name | Structure |
|---|---|---|
| Dopamine | Dopamine | 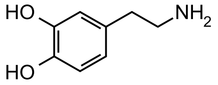 |
| LA/CA | Levodopa | 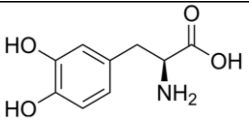 |
| | Carbidopa | 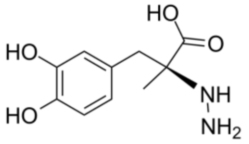 |
| | Pramipexole | 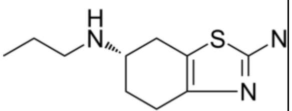 |
| | Ropinirole | 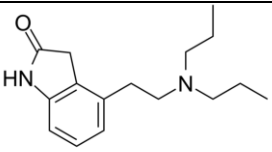 |
| | Rotigotine | 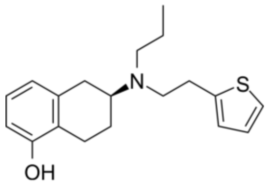 |
| | Bromocriptine | 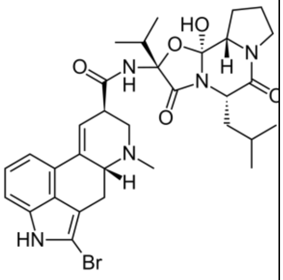 |
| | Pergolide | 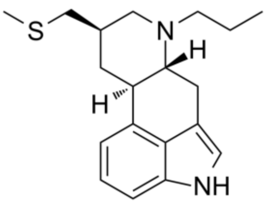 |
| | Cabergoline | 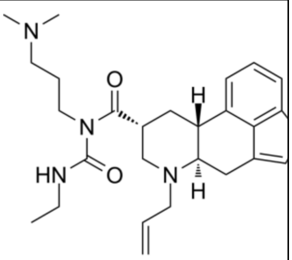 |
| Catechol-O-methyltransferase inhibitors | Tolcapone | 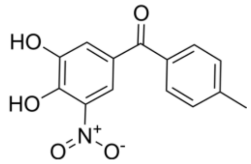 |
| | Entacapone | 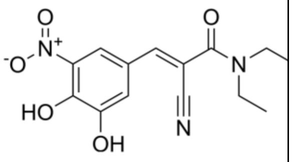 |
| Monoamine oxidase-B inhibitor | Rasagiline | 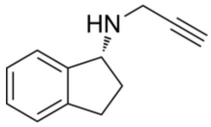 |

Table S2. Comparison between means of age and length of expanded allele of the groups in Table 1.

| | LA/CA | | | OTHER ANTI-PARKINSONIANS | | |
|---|---|---|---|---|---|---|
| | Controls | Cases | t (df), P value | Controls | Cases | t (df), P value |
| Age (years) | 52.5 ± 14.1 | 50.6 ± 14.2 | 0.98 (710), P=0.33 | 52.5 ± 14.2 | 54.0 ± 13.0 | 0.53 (652), P=0.60 |
| Expanded allele repeat length | 50.9 ± 24.0 | 57.3 ± 18.9 | 1.67 (443), P=0.10 | 50.7 ± 24.3 | 56.2 ± 17.8 | 0.95 (400), P=0.34 |

Table S3. Comparison between means of age and length of expanded allele of the groups in Table 3.

| | LA/CA | | | OTHER ANTI-PARKINSONIANS | | |
|---|---|---|---|---|---|---|
| | Controls | Cases | t (df), P value | Controls | Cases | t (df), P value |
| Age (years) | 53.2 ± 14.0 | 52.4 ± 12.5 | 0.35 (419), P=0.73 | 53.1 ± 14.1 | 55.1 ± 12.7 | 0.56 (378), P=0.58 |
| Expanded allele repeat length | 49.7 ± 23.6 | 56.2 ± 19.8 | 1.43 (276), P=0.16 | 49.4 ± 23.8 | 56.1 ± 19.7 | 0.92 (247), P=0.36 |

Table S4. Characteristics of the MJD/SCA3 group selected for the multivariate analysis of the effect of non-LA/CA anti-parkinsonian drugs.

| | Controls | Cases |
|---|---|---|
| n | 115 | 7 |
| >6 years monitoring, n* | 8 | 0 |
| Female, n (%) | 59 (51) | 2 (29) |
| Age (years)** | 51.3 ± 12.2 | 57.5 ± 15.1 |
| Expanded allele repeat length** | 71 ± 5.2 | 70 ± 4.7 |

*SARA scores monitored over periods longer than 6 years

**Mean ± standard deviation values

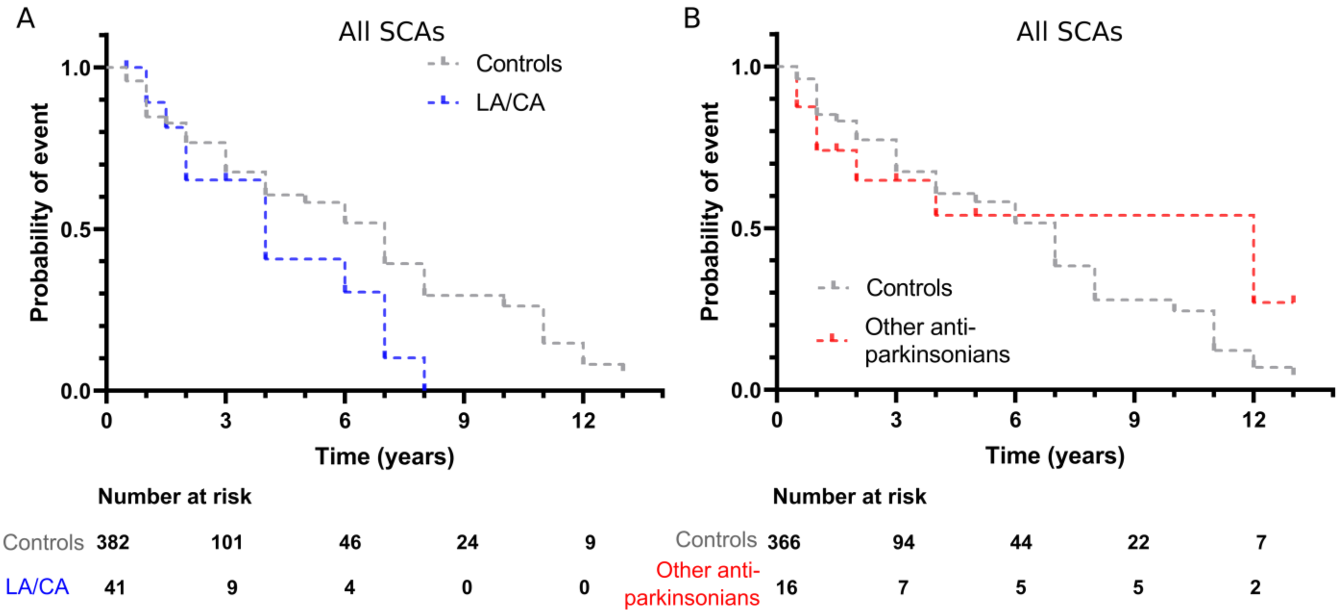


Figure S1. Kaplan-Meier analysis for events of a SARA score increase of > 4 points in spinocerebellar ataxia patients (A) who have previously taken LA/CA compared with controls, and (B) who have previously taken other anti-parkinsonian drugs compared with controls.

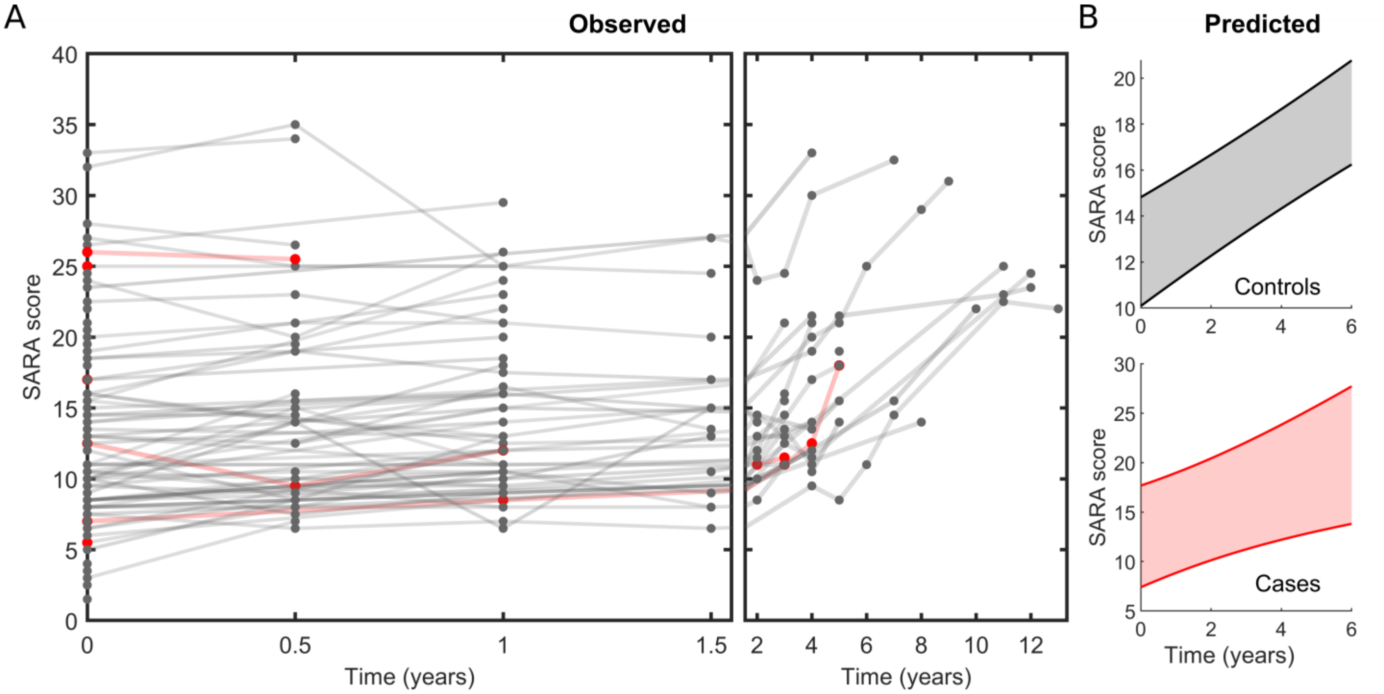


Figure S2. Effect of exposition to other anti-parkinsonian drugs on MJD/SCA3 evolution. Progress of SARA scores (A) observed in individuals with MJD/SCA3 and (B) predicted by the LME model for (top) controls and (bottom) cases of exposition to non-LA/CA anti-parkinsonian drugs. (B) 95% confidence intervals for the 6-year predictions computed assuming a random MJD/SCA3 patient with 58 years of age, and expanded allele length 70.

Table S5. Covariate-adjusted effect of exposition to non LA/CA anti-parkinsonian drugs on MJD/SCA3 progression. LME coefficients, 95% confidence intervals and P values fitted to the 13-year and 6-year SARA monitoring data.

| EFFECT | β | Lower | Upper | P value |
|---|---|---|---|---|
| **13 YEARS** | | | | |
| INTERCEPT | -34.54 | -56.42 | -12.66 | 0.002 |
| AGE | 0.21 | 0.11 | 0.32 | <0.001 |
| TIME | 1.01 | 0.82 | 1.19 | <0.001 |
| ALLELE | 0.52 | 0.26 | 0.77 | <0.001 |
| TREATMENT | 0.09 | -4.67 | 4.84 | 0.971 |
| TIME: TREATMENT | 0.32 | -0.69 | 1.34 | 0.530 |
| **6 YEARS** | | | | |
| INTERCEPT | -34.179 | -56.13 | -12.23 | 0.002 |
| AGE | 0.21071 | 0.10 | 0.32 | <0.001 |
| TIME | 1.0089 | 0.73 | 1.28 | <0.001 |
| ALLELE | 0.51241 | 0.26 | 0.77 | <0.001 |
| TREATMENT | 0.084112 | -4.68 | 4.84 | 0.972 |
| TIME: TREATMENT | 0.36096 | -0.61 | 1.33 | 0.465 |

Table S6. Characteristics of the MJD/SCA3 group selected for the multivariate analysis of the effect of anti-parkinsonian drugs.

| | Controls | Cases |
|---|---|---|
| n | 115 | 31 |
| >6 years monitoring, n* | 8 | 3 |
| Female, n (%) | 59 (51) | 17 (55) |
| Age (years)** | 51.3 ± 12.2 | 51.9 ± 14.0 |
| Expanded allele repeat length** | 71 ± 5.2 | 71 ± 5.7 |

*SARA scores monitored over periods longer than 6 years

**Mean ± standard deviation values

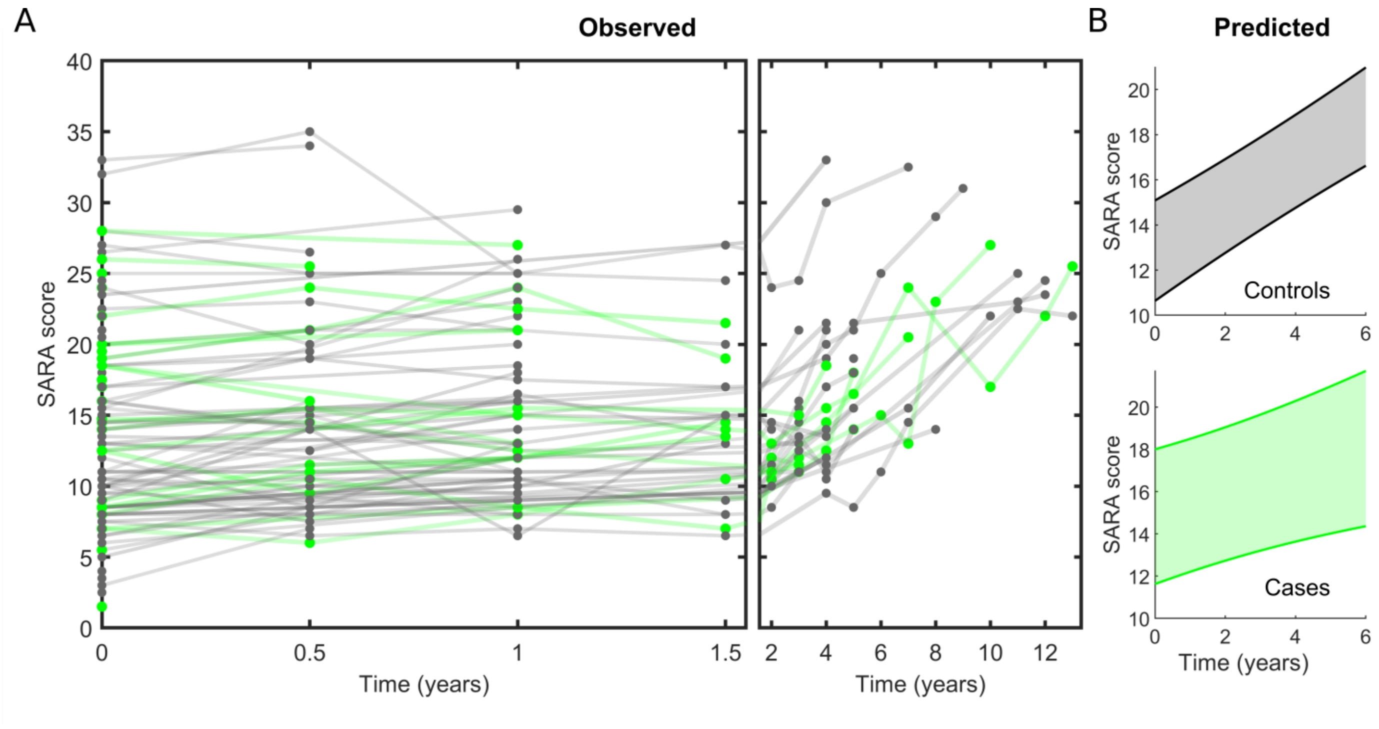


Figure S3. Effect of exposition to anti-parkinsonian drugs on MJD/SCA3 evolution. Progress of SARA scores (A) observed in individuals with MJD/SCA3 and (B) predicted by the LME model for (top) controls and (bottom) cases of exposition to anti-parkinsonian drugs. (B) 95% confidence intervals for the 6-year predictions computed assuming a random MJD/SCA3 patient with 58 years of age, and expanded allele length 70.

Table S7. Covariate-adjusted effect of exposition to anti-parkinsonian drugs on MJD/SCA3 progression. LME coefficients, 95% confidence intervals and P values fitted to the 13-year and 6-year SARA monitoring data.

| 13 YEARS | | | | |
|---|---|---|---|---|
| EFFECT | β | Lower | Upper | P value |
| INTERCEPT | -32.50 | -52.02 | -12.99 | 0.001 |
| AGE | 0.23 | 0.13 | 0.33 | <0.001 |
| TIME | 0.99 | 0.81 | 1.17 | <0.001 |
| ALLELE | 0.48 | 0.25 | 0.70 | <0.001 |
| TREATMENT | 1.82 | -0.53 | 4.17 | 0.129 |
| TIME: TREATMENT | -0.24 | -0.51 | 0.03 | 0.078 |
| 6 YEARS | | | | |
| INTERCEPT | -32.17 | -51.71 | -12.63 | 0.001 |
| AGE | 0.23 | 0.13 | 0.32 | <0.001 |
| TIME | 0.99 | 0.73 | 1.25 | <0.001 |
| ALLELE | 0.47 | 0.24 | 0.70 | <0.001 |
| TREATMENT | 1.96 | -0.41 | 4.32 | 0.105 |
| TIME: TREATMENT | -0.45 | -0.92 | 0.01 | 0.056 |

Table S8. Characteristics of the whole MJD/SCA3 group (early-symptomatic patients included) that was selected for the multivariate analysis of the CA/DA effect.

| | Controls | Cases |
|---|---|---|
| n | 131 | 24 |
| >6 years monitoring, n[*] | 9 | 3 |
| Female, n (%) | 64 (49) | 16 (67) |
| Age (years)[**] | 51.8 ± 12.4 | 50.7 ± 13.7 |
| Expanded allele repeat length[**] | 70.7 ± 5.2 | 71.7 ± 5.7 |

[*]SARA scores monitored over periods longer than 6 years

[**]Mean ± standard deviation values

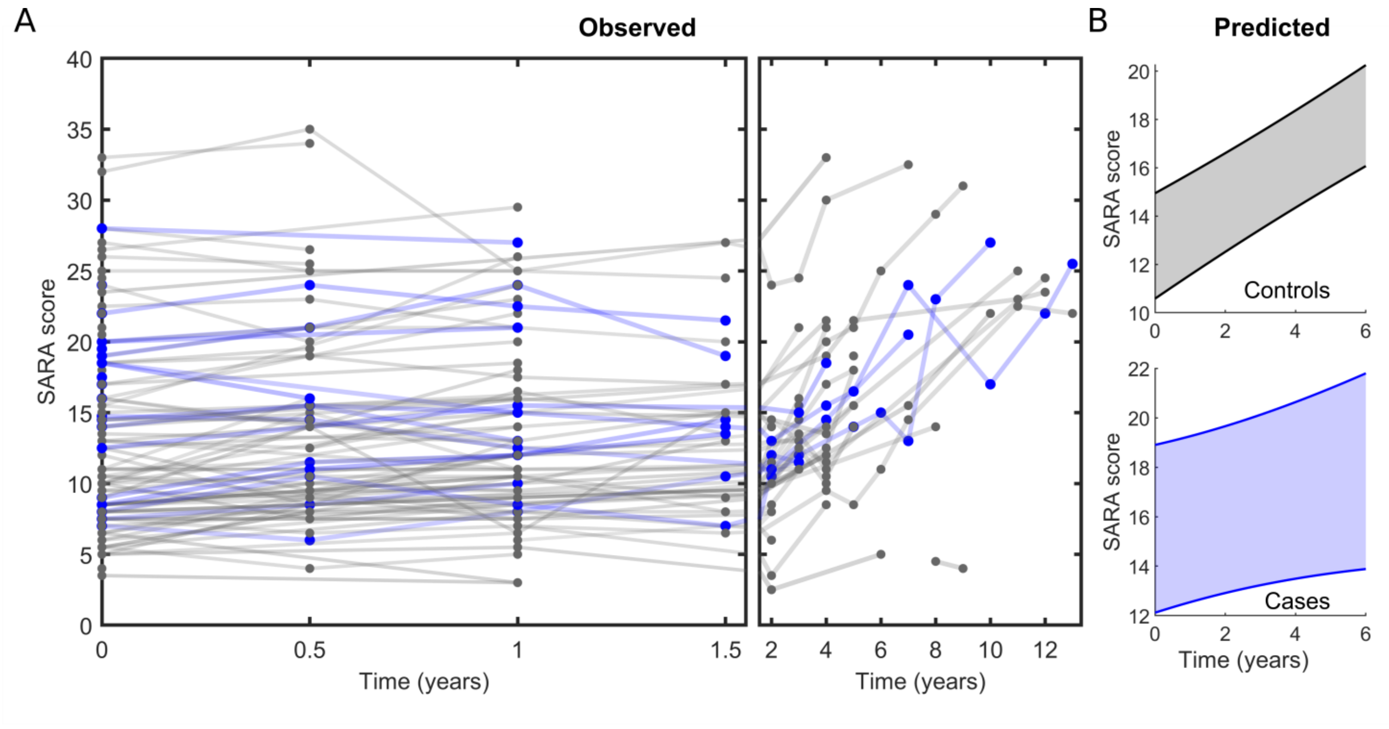


Figure S4. Effect of exposition to LA/CA on ataxia evolution of the whole MJD/SCA3 group (early-symptomatic patients included). Progress of SARA scores (A) observed in the whole group of MJD/SCA3 patients and (B) predicted by the LME model for (top) controls and (bottom) cases of exposition to LA/CA. (B) 95% confidence intervals for 6-year predictions computed assuming a random MJD/SCA3 patient with 58 years of age, and expanded allele length 70.

Table S9. Covariate-adjusted effect of exposition to LA/CA on ataxia evolution of the whole MJD/SCA3 group (early-symptomatic patients included). LME coefficients, 95% confidence intervals and P values fitted to the 13-year and 6-year SARA monitoring data.

| 13 YEARS | | | | |
|---|---|---|---|---|
| EFFECT | β | Lower | Upper | P value |
| INTERCEPT | -35.18 | -54.01 | -16.35 | <0.001 |
| AGE | 0.23 | 0.13 | 0.32 | <0.001 |
| TIME | 0.93 | 0.76 | 1.11 | <0.001 |
| ALLELE | 0.51 | 0.29 | 0.73 | <0.001 |
| TREATMENT | 2.66 | 0.03 | 5.29 | 0.048 |
| TIME: TREATMENT | -0.21 | -0.48 | 0.05 | 0.115 |
| 6 YEARS | | | | |
| INTERCEPT | -31.012 | -49.906 | -12.118 | 0.0014 |
| AGE | 0.208 | 0.113 | 0.304 | <0.001 |
| TIME | 0.899 | 0.661 | 1.137 | <0.001 |
| ALLELE | 0.465 | 0.247 | 0.683 | <0.001 |
| TREATMENT | 2.749 | 0.138 | 5.359 | 0.0391 |
| TIME: TREATMENT | -0.511 | -0.988 | -0.034 | 0.0357 |
